# Enrichment of patients with concomitant LATE on the Alzheimer’s disease continuum using hippocampal volume

**DOI:** 10.1101/2025.08.14.25333712

**Authors:** Nidhi S. Mundada, Xueying Lyu, Christopher A. Brown, Niyousha Sadeghpour, Emily McGrew, Long Xie, Philip A Cook, James Gee, Paul A. Yushkevich, Sandhitsu R. Das, David A. Wolk, the Alzheimer’s Disease Neuroimaging Initiative

## Abstract

**Introduction:** Clinical overlap between Alzheimer’s disease (AD) and limbic-predominant age-related TDP-43 encephalopathy (LATE), combined with absence of validated in vivo biomarkers, complicates identification of mixed AD/LATE pathology. We labeled individuals along the AD continuum with suspected concomitant LATE using the lower quartile hippocampal volume (HV) cut-off and examined associated atrophy and cognitive profiles.

**Methods:** We studied cognitively impaired ADNI participants with T1-MRI and amyloid- and tau-PET. Participants were classified into suspected (s) AD+sLATE-, AD-sLATE+, or AD+sLATE+ based on amyloid status and HV quartiles. Medial temporal lobe (MTL) and whole-brain atrophy patterns and cognitive profiles were compared cross-sectionally and longitudinally. Classification was validated in an autopsy cohort.

**Results:** AD+sLATE+ showed greater anterior hippocampal and amygdala atrophy than AD+sLATE-. AD-sLATE+ and AD+sLATE+ showed greater anterior MTL atrophy and worse memory and language performance. AD+sLATE+ also exhibited faster cognitive decline.

**Discussion:** A simple HV quartile cut-off may help identify mixed AD/LATE pathology and support clinical trial enrichment.

## 1. INTRODUCTION

Alzheimer’s disease (AD) is the leading cause of dementia, characterized by progressive cognitive decline driven by the accumulation of amyloid-beta (Aβ) plaques and tau neurofibrillary tangles.^1^ While Aβ deposition is an early hallmark of AD pathology, tau tangles, rather than amyloid plaques, are more strongly associated with neuronal loss and clinical progression, positioning tau as a primary driver of neurodegeneration in AD.^2–5^ However, many individuals with AD harbor additional age-related changes and neuropathologies that contribute to cognitive decline and neurodegeneration.^6–8^

One such co-pathology is limbic-predominant age-related TDP-43 encephalopathy (LATE), a neurodegenerative condition increasingly recognized as a major contributor to memory impairment in older adults.^9^ LATE neuropathic change (LATE-NC) primarily involves limbic structures such as the hippocampus and amygdala and is characterized by the pathological aggregation of TAR DNA-binding protein 43 (TDP-43).^10–12^ While its clinical presentation often resembles that of AD, LATE-NC follows a distinct progression trajectory and frequently coexists with AD pathology, complicating differential diagnosis. The high prevalence of LATE among individuals with AD—estimated to be present in up to 50% of individuals with advanced AD (Braak stage V–VI) in community-based autopsy cohorts^13^—limits the ability to attribute observed atrophy and clinical symptoms solely to AD pathology, given that standard biomarkers capture amyloid and tau, but do not account for TDP-43 pathology. Furthermore, individuals with both AD and LATE pathologies tend to experience faster cognitive decline and greater neurodegeneration than those with AD alone.^14–16^ This raises additional questions about whether the presence of LATE-NC copathology modulates response to AD-targeted treatments, such as anti-amyloid immunotherapy.^17^ These uncertainties underscore the need for better stratification methods to identify individuals with likely LATE co-pathology, both to improve diagnostic and prognostic precision as well as to guide selection of clinical treatment and trial enrollment.

Identifying LATE during life remains challenging due to the absence of clinically available PET or biofluid biomarkers for TDP-43 pathology. While ongoing research efforts are developing promising candidates, these are not yet ready for clinical implementation.^18,19^ Generally, investigators have examined indirect markers of LATE-NC—including both structural^20,21^ and metabolic^22–24^ neuroimaging—to differentiate LATE from AD. While hippocampal atrophy is a hallmark of both AD neuropathological change (ADNC) and LATE-NC, Yu et al.^25^ demonstrated that autopsy-confirmed cases with both ADNC and LATE-NC or LATE-NC alone, exhibited significantly smaller postmortem hippocampal volumes than those with ADNC alone. Notably, among individuals with ADNC, those in the lower range (∼lowest quartile) of hippocampal volume appeared disproportionately enriched for concomitant LATE-NC, suggesting that TDP- 43 contributes to the variability in atrophy severity within AD. Together, these findings indicate that TDP-43 pathology contributes to neurodegeneration above and beyond what is explained by amyloid and tau burden alone, and may account for observed heterogeneity within the AD continuum. Indeed, a recent expert consensus group proposed clinical criteria for diagnosing possible or probable LATE, emphasizing not only the presence of memory impairment, but structural MRI evidence of substantial hippocampal atrophy.^26^

While prior studies have proposed MRI-based predictors of LATE-NC^20,21,27,28^, these approaches often involve complex multivariate models that may be difficult to implement in clinical settings. In contrast, we tested a simple, quartile-based approach using hippocampal volume to identify individuals who may harbor LATE-NC within the AD continuum. We derived hippocampal volume quartiles across the AD continuum and defined the lowest quartile as atrophy beyond what is “typically” observed in AD, which we expect to be due to, in part, underlying LATE co-pathology. Furthermore, we hypothesized that these individuals would exhibit distinct patterns of neurodegeneration and atrophy on structural MRI that have previously been associated with LATE-NC (e.g. a greater anterior-to-posterior gradient of medial temporal lobe atrophy^21,29,30^), as well as other clinical features suggestive of AD with concomitant LATE-NC, such as accelerated cognitive decline.^14^ To assess the pathological validity of this approach, we further evaluated it in an independent autopsy cohort. By refining identification of individuals along the AD continuum—particularly those with concomitant LATE copathology—this study aims to advance our understanding of neurodegenerative heterogeneity. This approach has the potential to enhance cohort selection in research, refine biomarker studies, and contribute to more personalized strategies for diagnosing and treating mixed etiology dementia.

## 2. METHODS

### 2.1. Participants

#### 2.1.1 Main cohort

Participants were retrospectively selected from the Alzheimer’s Disease Neuroimaging Initiative (ADNI) study (http://adni.loni.usc.edu). We applied a hippocampal volume cut-off to cognitively impaired (CI) individuals who met the following criteria: 1) clinical diagnosis of mild cognitive impairment (MCI) or dementia, and 2) amyloid-PET, tau-PET and 3 Tesla T1-weighted MRI scan within 365 days of each other. Amyloid positivity was determined based on amyloid-PET: 232 individuals were classified as amyloid-positive (Aβ+), and 165 individuals were classified as amyloid-negative (Aβ-). Enrichment criteria were applied to the total sample, refer to section 2.3 on details of stratification and Supplementary Figure 1 for cohort selection flowchart. In addition, we included Aβ- cognitively unimpaired (CU) individuals who were matched to the CI group on age, sex, and education and had complete imaging data, to serve as a reference group.

#### 2.1.2. Autopsy cohort

ADNI participants with neuropathologic assessments and T1-weighted MRI, either 1.5 Tesla or 3 Tesla, were included as a validation cohort. Using neuropathologic assessments, we characterized individuals with a definitive diagnosis using their Alzheimer’s disease neuropathologic change (ADNC) assessments and presence of TDP-43 in their hippocampus.

Individuals with intermediate/high ADNC and no hippocampal TDP-43 were classified as AD+LATE- (n=44), and those with intermediate/high ADNC and hippocampal TDP-43 as AD+LATE+ (n=35). Of the 79 participants, 69 were CI at the time of their baseline scan, while 10 were CU, 8 of whom converted to CI prior to death.

### 2.2. Image acquisition and processing

#### 2.2.1. Image acquisition

Individuals in the main analysis had 3 Tesla T1-weighted MRI scans of resolution 1.0×1.0×1.2 mm^3^ or 1.0×1.0×1.0 mm^3^ while those in the autopsy sample had either 1.5 Tesla T1-weighted MRI (n=39) of resolution 1.25×1.25×1.2 mm^3^ or a 3 Tesla T1-weighted MRI (n=40) of resolution 1.0×1.0×1.2 mm^3^ or 1.0×1.0×1.0 mm^3^.

Amyloid-PET imaging was acquired using either ^18^F-florbetapir (50-70 minutes post-injection) or ^18^F-florbetaben (90-110 minutes post-injection). Tau-PET imaging was performed using six 5- minute frames acquired between 75-105 minutes post-injection ofL^18^F-Flortaucipir.

Preprocessed tau-PET images with a uniform 6 mm full-width-at-half-maximum (FWHM) resolution were obtained from the ADNI archive (“Coreg, Avg, Std Img and Vox Size, Uniform Resolution”).

#### 2.2.2. T1-MRI processing

The Automatic Segmentation of Hippocampal Subfields-T1 (ASHS-T1, https://sites.google.com/view/ashs-dox/) pipeline was used to automatically segment the MTL subregions.^31^ Segmentations were performed on each participant’s T1-MRI scan closest in time to their tau-PET scan (used for cross-sectional analyses), as well as all available T1-MRIs within ±5 years from the scan used for cross-sectional analyses. The segmented subregions included the anterior and posterior hippocampus (AH/PH), amygdala, entorhinal cortex (ERC), Brodmann areas 35 and 36 (BA35, BA36), and the parahippocampal cortex (PHC). ASHS-T1 is specifically designed to address confounds like inclusion of dura mater, which limit traditional whole-brain segmentation methods.^32^ For each subregion, a summary region of interest (ROI) metric was extracted: volumetric measurements were computed for AH, PH and amygdala, while median cortical thickness was derived for the MTL cortical regions (ERC, BA35, BA36, and PHC) using the “cortical reconstruction for ASHS (CRASHS)” surface-based pipeline applied to the ASHS- T1 output. Details of the CRASHS pipeline have been outlined here.^33^ Briefly, CRASHS applies surface-based cortical modeling and diffeomorphic registration to ASHS-T1 segmentations, enabling the extraction of median cortical thickness for MTL cortical regions, as well as pointwise thickness maps with consistent anatomical correspondence across participants enabling vertex-wise statistical analyses of regional disease effects. Of the 2554 thickness maps from the main analysis, left and right separate, from 386 participants, including cross-sectional and longitudinal timepoints, 0.7% failed processing. Intracranial volume (ICV) was estimated from each participant’s structural MRI using in-house segmentation software built on the ASHS framework.

Additionally, T1w-MRI scans were bias-corrected and skull-stripped using Advanced Normalization Tools (ANTs), followed by cerebellar, cortical, and subcortical parcellation via multi-atlas Joint Label Fusion with the BrainColor atlas, which defines 102 ROIs.^34,35^ Whole brain thickness maps were generated using the DiReCT cortical thickness estimation method.^36^

For longitudinal analyses, T1w scans were processed using a longitudinal ANTs-based pipeline that incorporates symmetric diffeomorphic registration to generate within-subject templates and ensures consistent cortical thickness estimation across timepoints, as validated in prior work.^37^

For longitudinal analysis, we included scans ±5 years from the timepoint used in the cross-sectional analyses. Among 386 participants, 304 had longitudinal MRI scans, with an average of 3.9±2.1 scans per participant over a follow-up span of 0.5-9.8 years.

#### 2.2.3. PET processing

Rigid-body registration of tau-PET to T1-weighted MRI was performed using ANTs,^38^ and all registrations were visually inspected for quality assurance. Standardized uptake value ratio (SUVR) maps were computed using an inferior cerebellar reference region, and mean SUVR values were extracted from all cortical and subcortical ROIs following partial volume correction.

To quantify tau burden, Tau-MaX, a combined measure of tau extent and magnitude, was computed using a Gaussian mixture modeling approach to distinguish pathologic from non-pathologic signal from tau-PET scans. Details on computing Tau-MaX have been previously described.^39^ Tau-MaX values were calculated for both a global cortical region set and a temporal meta-ROI, yielding global and temporal specific Tau-MaX measures used in subsequent analyses.

Amyloid positivity was determined using amyloid-PET SUVRs provided by ADNI PET Core. Patients were classified as Aβ+ based on predetermined thresholds:^40^ SUVR ≥ 1.11 for ^18^F-Florbetapir and 1.08 for ^18^F-Florbetaben.

### 2.3. Cohort stratification: Percentile-based grouping

We assessed the distribution of total hippocampal volume (AH+PH) in patients with MCI or dementia due to AD. We used the total hippocampal volume of the most affected hemisphere, i.e., the smaller of the two hippocampal volumes, rather than taking a mean of the two hemispheres, as LATE-NC is frequently asymmetric.^10^ Age at MRI and ICV-adjusted hippocampal volume was calculated for all patients using the regression estimates from all CU individuals; sex was not included as a covariate since ICV accounted for sex differences.

Using a quartile-based approach, we classified Aβ+ patients into suspected (s) “AD+sLATE-” and “AD+sLATE+” groups based on the distribution of adjusted hippocampal volume among Aβ+ patients. Patients in the first quartile (Q1, percentile < 25) were classified as “AD+sLATE+”, and patients in the 3^rd^ and 4^th^ quartile (Q3-Q4, percentile > 50) were classified as “AD+sLATE-”. Patients in the second quartile (Q2, 25 < percentile < 50) were excluded.

We included two Aβ- reference groups, Aβ-CI and Aβ-CU. The same cut-off from the Aβ+ patients was applied to the Aβ-CI participants to select patients with significantly low hippocampal volume (Q1). Given this level of hippocampal atrophy, these patients were suspected to have LATE-NC as the primary underlying pathology, identified as “AD-sLATE+” in this study. The Aβ-CU group served as an additional reference group.

In summary, we had four groups, two disease of interest groups defined using the quartile approach for hippocampal volume, 1) AD+sLATE- (Aβ+CI, Q3-Q4), 2) AD+sLATE+ (Aβ+CI, Q1), and two reference groups, 3) AD-sLATE+ (Aβ-CI, Q1), and 4) Aβ-CU.

### 2.4. Statistical analysis

Statistical analyses were performed using R version 4.4.2 (www.r-project.org), except for cross-sectional pointwise MTL thickness analyses, which were carried out in Python with the meshglm tool from the CM-Rep package (github.com/pyushkevich/cmrep) and cross-sectional voxelwise whole brain analyses, which were carried out using FSL tools.^41^ All tests were two-sided.

#### 2.4.1. Region level analyses and imaging features

We assessed group differences in ROI volumes (AH, PH, and amygdala) and derived imaging features using pairwise t-tests. Imaging features were selected based on a priori research suggestive of the presence of LATE-NC. Ratio of ERC/PHC thickness was computed, where lower ratios suggest more thinning in the anterior MTL. Since LATE is often asymmetric, we calculated hippocampal asymmetry index as follows: abs((L-R)/(L+R)*200), where higher values indicate greater asymmetry between the two hemispheres; all values are positive since we take the absolute value.

#### 2.4.2. MTL pointwise analyses

For MTL pointwise group comparisons, general linear model (GLM) testing was conducted at each vertex of the cortical thickness maps. Permutation testing using the threshold-free cluster enhancement (TFCE) approach^42^ with parameters EL=L0.5, HL=L2, and ΔhL=L0.01 and 10,000 permutations was used to compute family-wise error rate (FWER)-corrected p-values at each vertex. Age was used as a covariate when comparing AD+sLATE-, AD-sLATE+, and AD+sLATE+ groups to Aβ-CU; age and temporal Tau-MaX were used as covariates when comparing AD+sLATE- vs AD+sLATE+ groups to control for the effect of tau pathology, a surrogate for AD progression, for cross-sectional analyses. For longitudinal analyses, we fit a linear-mixed effects (LME)^43^ model at every vertex with random intercepts with false discovery rate (FDR) correction. Covariates included are in the model equations below:

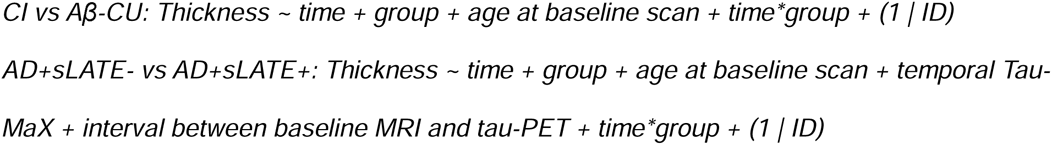

#### 2.4.3. Whole brain analyses

For whole brain voxelwise cross-sectional analyses, cortical thickness maps were analyzed by using the TFCE method with parameters E=0.5, H=2, and Δh=0.01 and 10000 permutations using FSL Randomise. Age was used as a covariate for AD+sLATE-, AD-sLATE+, and AD+sLATE+ vs Aβ-CU group comparisons; age and global Tau-MaX were used for AD+sLATE- vs AD+sLATE+ group comparisons. For longitudinal analyses, we fit an LME model at every voxel with random intercepts using the lmerNIfTI package in R^44^ and used the same equations as above, except global Tau-MaX was used instead of temporal Tau-MaX. For both cross-sectional and longitudinal analyses, comparisons between CI and Aβ-CU groups are shown at p<0.01 FWER- or FDR-corrected to highlight only the most robust effects, as a p<0.05 threshold yielded widespread significance. Comparisons between patient groups are shown at p < 0.05 FWER- or FDR-corrected to retain sensitivity to more subtle group differences.

#### 2.4.4. Cognitive domain analyses

We evaluated group differences across three cognitive domains: memory, executive function, and language using ADSP-PHC composite scores provided by ADNI. Pairwise t-tests were performed to assess group differences and analysis of variance was performed when controlling for global Tau-MaX in the models. For cross-sectional analysis, we used the psychometric assessment closest to the MRI scan used in the imaging cross-sectional analyses above. For longitudinal analyses, we included all available assessments ±5 years from the timepoint used in the cross-sectional analyses. LME models with random slopes and random intercepts were used. To assess differences in rates of decline between groups, we performed pairwise t-tests on the interaction between time*group from the LME models. Covariates included are in the model equation below:

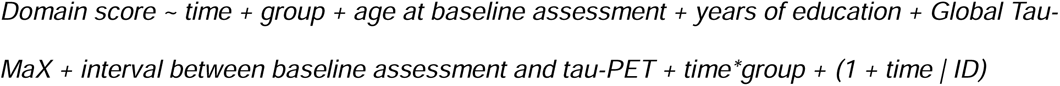

#### 2.4.5. SILA analysis of cognitive trajectories

We applied the sampled iterative local approximation (SILA) algorithm^45^ to estimate the distribution of the age at which individuals become cognitively impaired relative to the age of tau onset. To estimate age of tau onset, we applied the SILA model using Global Tau-MaX values from our previously described dataset^39^ following the approach outlined by Betthauser et al.^45^ Briefly, SILA estimates a biomarker trajectory by first using discrete sampling to quantify the rate of change at each biomarker level, followed by robust LOESS smoothing and numerical integration via Euler’s method. Once a positivity threshold is defined, the resulting trajectory can be aligned to the time of biomarker positivity, allowing estimation of time from positivity for any given biomarker value. Tau positivity was defined using the 97.5^th^ percentile of global Tau-MaX values in Aβ-CU individuals, yielding a cut-off of >3.31 and tau onset age was estimated based on the final event point with an extrapolation of 3 years and truncation of estimated ages to the ages observed in the original model. Then, for each patient group, we fitted separate SILA models to cognitive measures to evaluate differences in cognitive trajectories. For these models, we anchored cognitive models to tau age (rather than chronological age) using estimated tau onset age. We then estimated that age of cognitive impairment onset using the group-specific model final event point and extrapolation of 5 years and truncation of estimated ages to the ages observed in the original model. The threshold for cognitive impairment for each domain was defined using a threshold of 1.5 SD below the mean of CU participants and scaled by -1 as SILA associates higher values with higher disease severity.

#### 2.4.6. Validation in Autopsy-confirmed cohort

Finally, we evaluated the quartile-based enrichment strategy in an autopsy-confirmed sample from ADNI. Hippocampal volume cutoff derived from Aβ+ patients in the main cohort was applied to categorize patients in the first quartile (severe hippocampal atrophy) and third/fourth quartile (less severe hippocampal atrophy). Within each quartile-based group, we assessed the frequency of ADNC with and without LATE-NC from the neuropathologic assessments available in ADNI. Pointwise MTL thickness differences across AD+LATE- and AD+LATE+ groups were also assessed using GLM, controlling for age at scan, Braak stage, antemortem interval (AMI) and field strength.

## 3. RESULTS

Applying a quartile-based cut-off on adjusted hippocampal volumes in CI individuals from ADNI yielded 115 individuals with AD+sLATE- (Aβ+, Q3-Q4, volume > 2861.4 mm^3^), 58 with AD+sLATE+ (Aβ+, Q1, volume < 2502.2 mm^3^) and 20 with AD-sLATE+ (Aβ-, Q1). Summary characteristics are reported in Table 1. As expected by definition of the groups, AD+sLATE+ had significantly lower hippocampal volumes compared to AD+sLATE- but did not differ from AD- sLATE+. AD+sLATE- and AD-sLATE+ had lower tau burden and load, as measured by temporal and global Tau-MaX, compared to AD+sLATE+, suggesting some degree of greater AD progression in the AD+sLATE+ group than in the AD+sLATE- group.

**Table 1:**
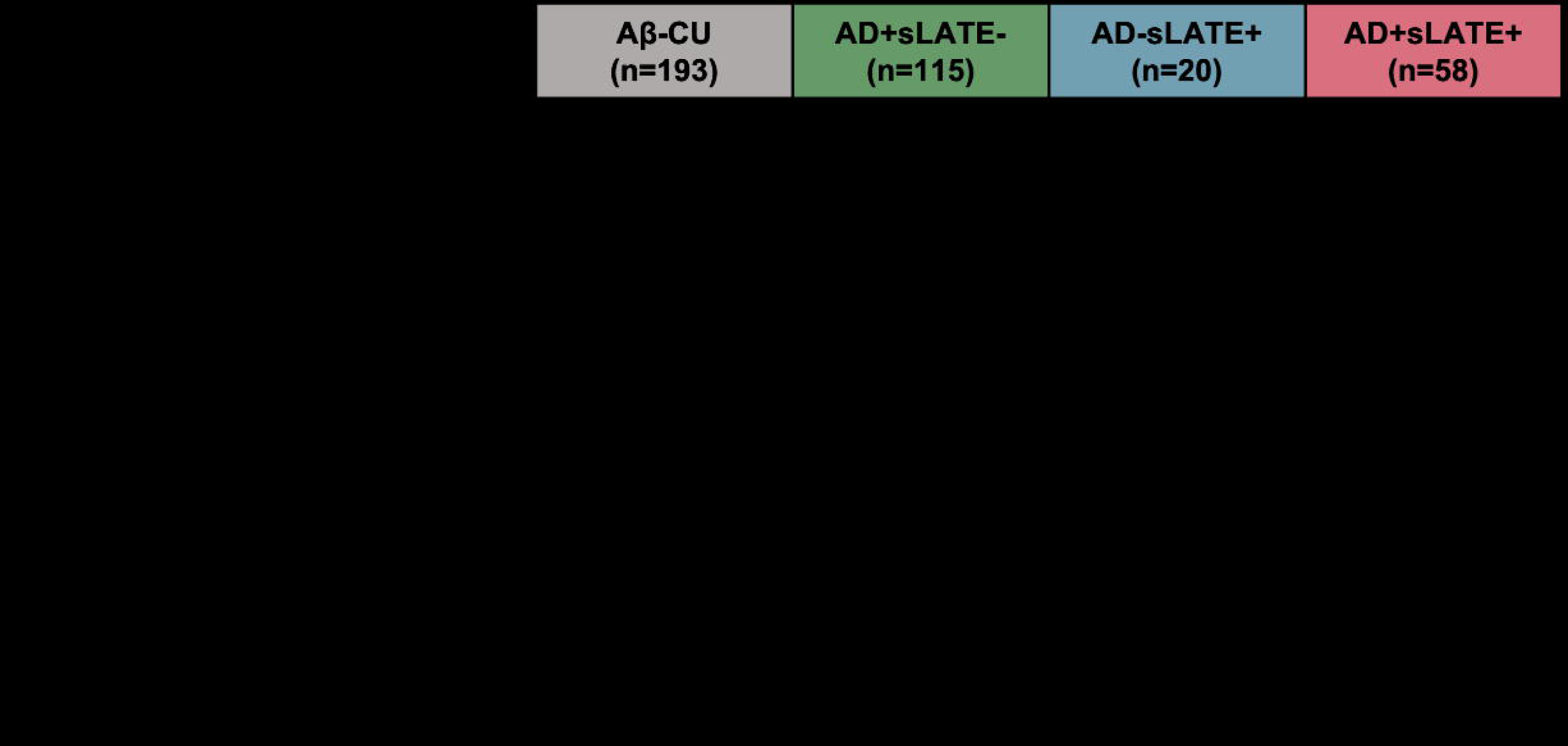
Demographics and Cohort Characteristics. Pairwise group comparisons are reported for AD+sLATE- vs AD+sLATE+ (asterisk in AD+sLATE- column) and AD-sLATE+ vs AD+sLATE+ group (asterisk in AD-sLATE+ column) - significance levels are indicated as follows: p ≤ 0.05 (*), p ≤ 0.001 (***). Hippocampal volume is adjusted for age and intracranial volume.

### 3.1. Regional-level analysis and imaging features

Plots in Figure 1A-C display that AD+sLATE- had lower mean posterior hippocampus volume, but not anterior hippocampus volume compared to Aβ-CU, while the AD+sLATE+ showed lower anterior and posterior hippocampus as well as amygdala volumes, possibly indicating greater anterior involvement of TDP-43 although AD+sLATE- displayed a significant amygdala difference, albeit significantly less so than the sLATE+ groups. Despite AD+sLATE- and AD+sLATE+ differing in tau load, AD+sLATE- covered a wide range of global Tau-MaX with significant overlap with the AD+sLATE+ group (Figure 1D).

**Figure 1.**
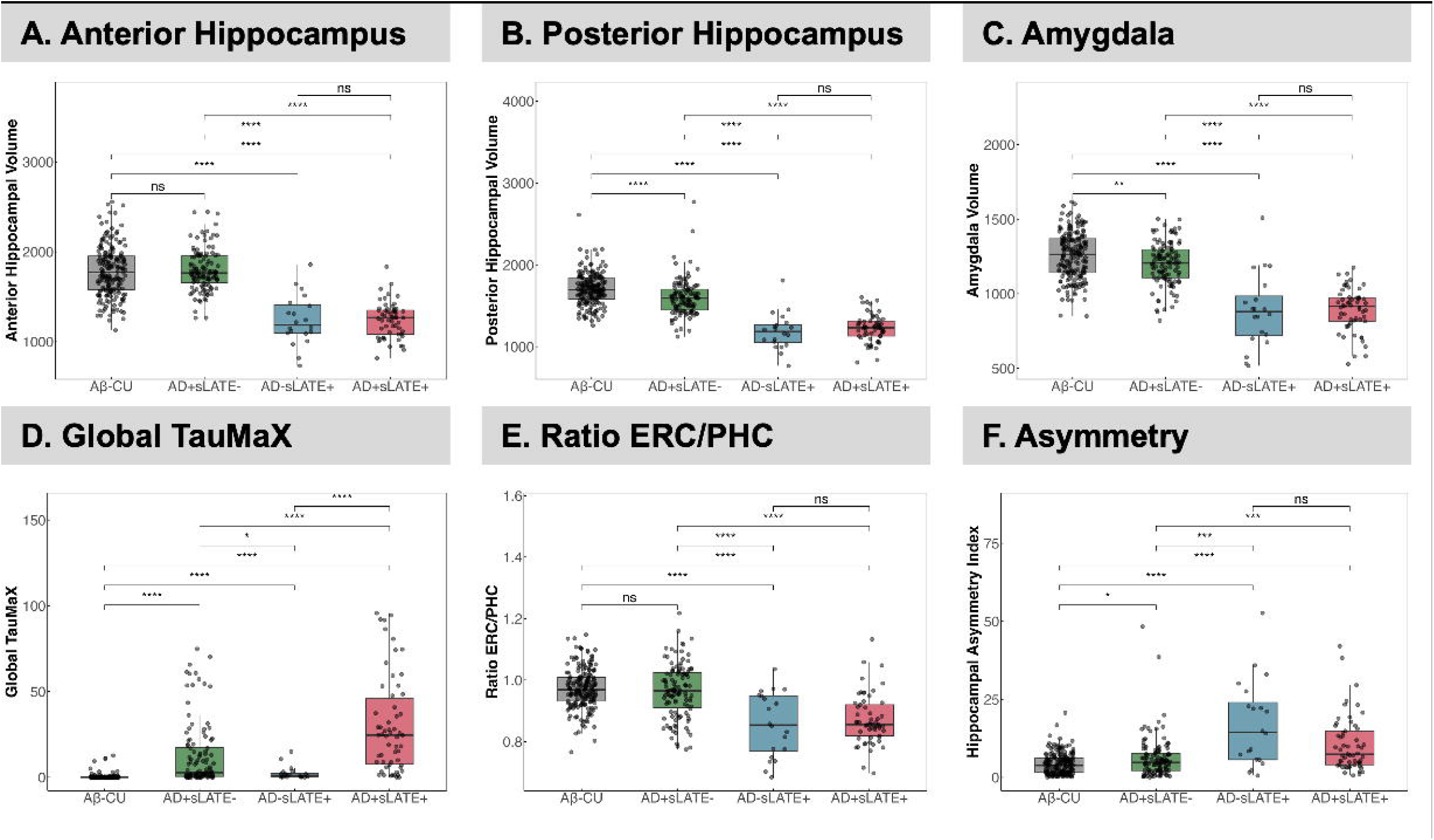
Differences between ROIs and imaging features across suspected groups. **A-D.** Distribution of anterior/posterior hippocampal volume, amygdala volume and tau burden as measured by Global Tau-MaX from tau-PET across groups. **E-F.** Distribution of additional features suggestive of LATE, from prior work, ratio of ERC to PHC, and hippocampal asymmetry calculated as (|(L-R)| / (L+R))*200. Significance levels are indicated as follows: p ≤ 0.05 (*), p ≤ 0.01 (**), p ≤ 0.001 (***), and p ≤ 0.0001 (****). ’ns’ indicates not significant (p > 0.05).

Our previous work has shown that lower ERC/PHC ratio can discriminate TDP-43 positive individuals with intermediate/high ADNC from those without TDP-43 with AUC of 0.82.^21^ Here, we found that AD+sLATE+ showed lower ERC/PHC ratio, similar to AD-sLATE+ (Figure 1E). Studies have shown that individuals with LATE also tend to show asymmetric atrophy,^10^ and we found that both AD+sLATE+ and AD-sLATE+ groups had greater asymmetry indices compared to AD+sLATE- and Aβ-CU groups (Figure 1F).

### 3.2. Pointwise analysis of medial temporal lobe thickness

#### 3.2.1. Cross-sectional

Compared to Aβ-CU, AD+sLATE- showed lower thickness in the anterior ERC and BA35 when controlling for age; however, AD-sLATE+ and AD+sLATE+ showed stronger effects in these regions with a clear anterior-to-posterior gradient in the MTL cortex (Figure 2A I-III). Controlling for AD progression with temporal Tau-MaX, the AD+sLATE+ group displayed lower thickness in the anterior regions of the MTL compared to the AD+sLATE- group (Figure 2A IV).

**Figure 2:**
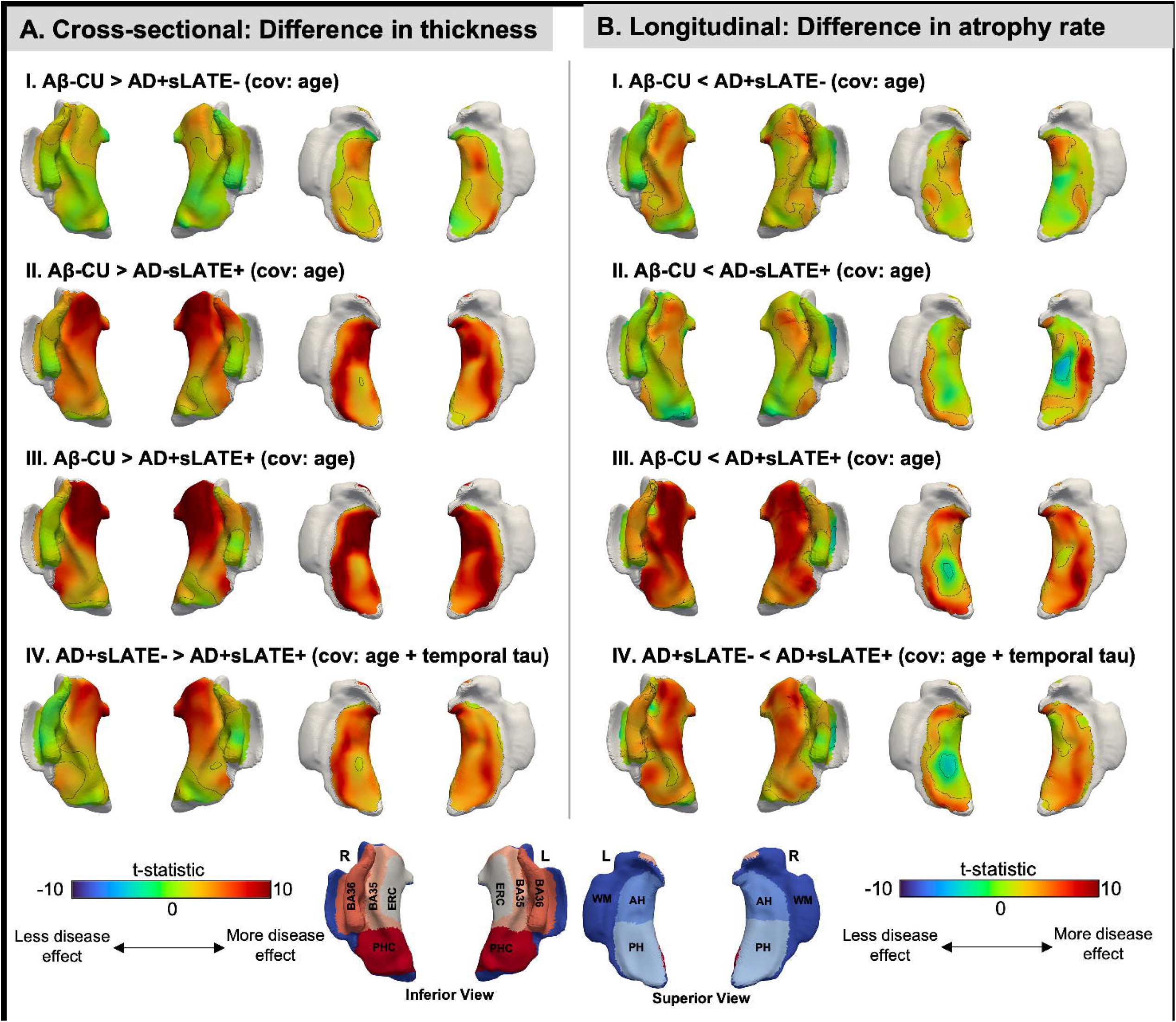
Pointwise structural cross-sectional and longitudinal group differences in the MTL. **A.** Maps represent group differences in thickness obtained using a surface-based MTL analysis pipeline, CRASHS. Each 3D model represents the inflated representation of the mid-surface between the MTL gray matter and white matter, shown either from superior or inferior. Age (I-IV) and temporal Tau-MaX (IV) were used as covariates. Tau-MaX is a measure which accounts for both the amount and extent of tau burden. Black outlines represent clusters significant at p<0.05 FWER-corrected (using the threshold-free cluster enhancement (TFCE) method with 10,000 permutations). **B.** Linear mixed-effects models were run at every point in the MTL thickness maps. For comparisons with CU (I-III), the following equation was used: Thickness ∼ time + group + age at baseline scan + time*group + (1 | ID). For comparisons between AD groups (IV), the following equation was used: Thickness ∼ time + group + age at baseline scan + temporal Tau-MaX + interval between baseline MRI and tau PET + time*group + (1 | ID). Black outlines represent clusters significant at p<0.05 FDR-corrected (using false discovery rate). AH=anterior hippocampus, PH=posterior hippocampus, ERC=entorhinal cortex, BA35/36=brodmann areas 35/36, PHC=parahippocampal cortex, WM=white matter.

#### 3.2.2. Longitudinal

Compared to Aβ-CU, AD+sLATE- showed limited atrophy throughout the MTL cortex, while AD- sLATE+ showed atrophy confined to the ERC and BA35; AD+sLATE+ showed severe atrophy throughout the MTL cortex, as well as the hippocampus (Figure 2B I-III). Additionally, when comparing AD+sLATE- to AD+sLATE+, controlling for temporal Tau-MaX at baseline, AD+sLATE+ displayed significantly greater rate of atrophy, particularly within cortical MTL regions (Figure 2B IV).

### 3.3. Voxelwise analysis of whole brain thickness

#### 3.3.1. Cross-sectional

Compared to Aβ-CU, AD+sLATE- showed lower thickness in the temporal and parietal regions consistent with typical AD. AD-sLATE+ showed lower thickness largely confined to the medial and anterior temporal and fronto-insular regions. AD+sLATE+ showed lower thickness throughout the cortex (Figure 3A I-III). Interestingly, when comparing AD+sLATE- and AD+sLATE+ and controlling for global Tau-MaX to account for AD severity, AD+sLATE+ displayed lower thickness almost exclusively in the medial and anterior temporal lobe regions at p<0.05 FDR-corrected threshold similar to that of AD-sLATE+ (Figure 3A IV).

**Figure 3:**
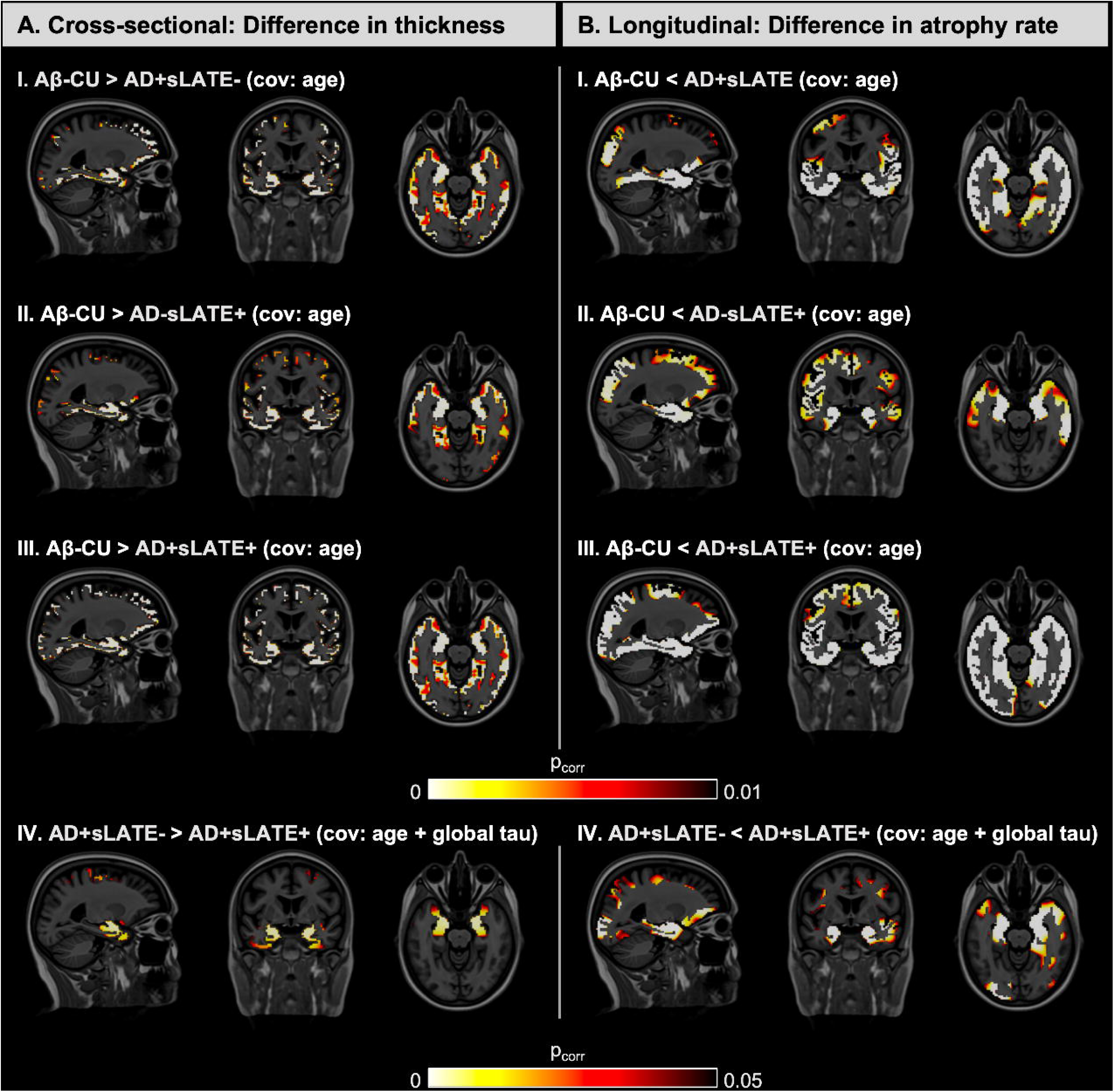
Voxelwise structural cross-sectional and longitudinal group differences in the whole brain. **A.** Voxelwise group comparisons in whole brain thickness maps for controls vs patient groups (I-III) are shown at p<0.01 FWER-corrected, and between patient groups (IV) are shown at p<0.05 FWER-corrected. Age (I-IV) and Global Tau-MaX (IV) at baseline were used as covariates. Tau-MaX is a measure which accounts for both the amount and extent of tau burden. **B.** Linear mixed-effects models were run at every voxel in the whole brain thickness maps. For comparisons with CU (I-III), the following equation was used: Thickness ∼ time + group + age at baseline scan + time*group + (1 | ID). For comparisons between AD groups (IV), the following equation was used: Thickness ∼ time + group + age at baseline scan + Global Tau-MaX + interval between baseline MRI and tau PET + time*group + (1 | ID). P-value of the interaction between group*time for comparisons with CU (I-III) are shown at p<0.01 FDR-corrected, and between AD groups (IV) are shown at p<0.05 FDR-corrected.

#### 3.3.2 Longitudinal

Results were generally consistent with cross-sectional findings, though they revealed a more extensive pattern of significant effects. When compared to Aβ-CU, AD+sLATE- displayed a significantly higher rate of atrophy in the temporal and parietal regions, AD-sLATE+ in the temporal lobe, and AD+sLATE+ throughout the cortex (Figure 3B I-III). Comparison between AD+sLATE- and AD+sLATE+ when controlling for Tau-MaX and for the interval between tau- PET and the baseline MRI scan in addition to age, again revealed that the AD+sLATE+ group had a greater rate of atrophy than AD alone most prominently and largely confined to medial and anterior temporal lobe regions, as well as orbitofrontal (Figure 3B IV). Nonetheless, there was some evidence of posterior cortical involvement as well.

### 3.4. Differences between groups across cognitive domains

#### 3.4.1. Cross-sectional

Groups differed from each other in composite memory score. AD+sLATE+ had the lowest score, followed by AD-sLATE+, AD+sLATE- and then Aβ-CU. Furthermore, when controlling for global tau, AD+sLATE+ differed from AD+sLATE- and AD-sLATE+, suggesting memory impairment beyond the effect of tau-related neurodegeneration. Pairwise comparisons revealed significant differences in executive function between all groups. However, AD+sLATE+ no longer significantly differed from AD+sLATE- or AD-sLATE+ when controlling for global tau, likely indicating minimal effect of non-tau related neurodegeneration on executive function. Finally, all groups, except AD-sLATE+ vs AD+sLATE+, also showed significant differences in language.

However, when controlling for tau, the difference between AD+sLATE- vs AD+sLATE+ was no longer significant (Figure 4A). Thus, this suggests that once the effects of AD severity based on tau are accounted for, the only remaining cross-sectional difference is in memory, which is primarily affected by LATE.

**Figure 4.**
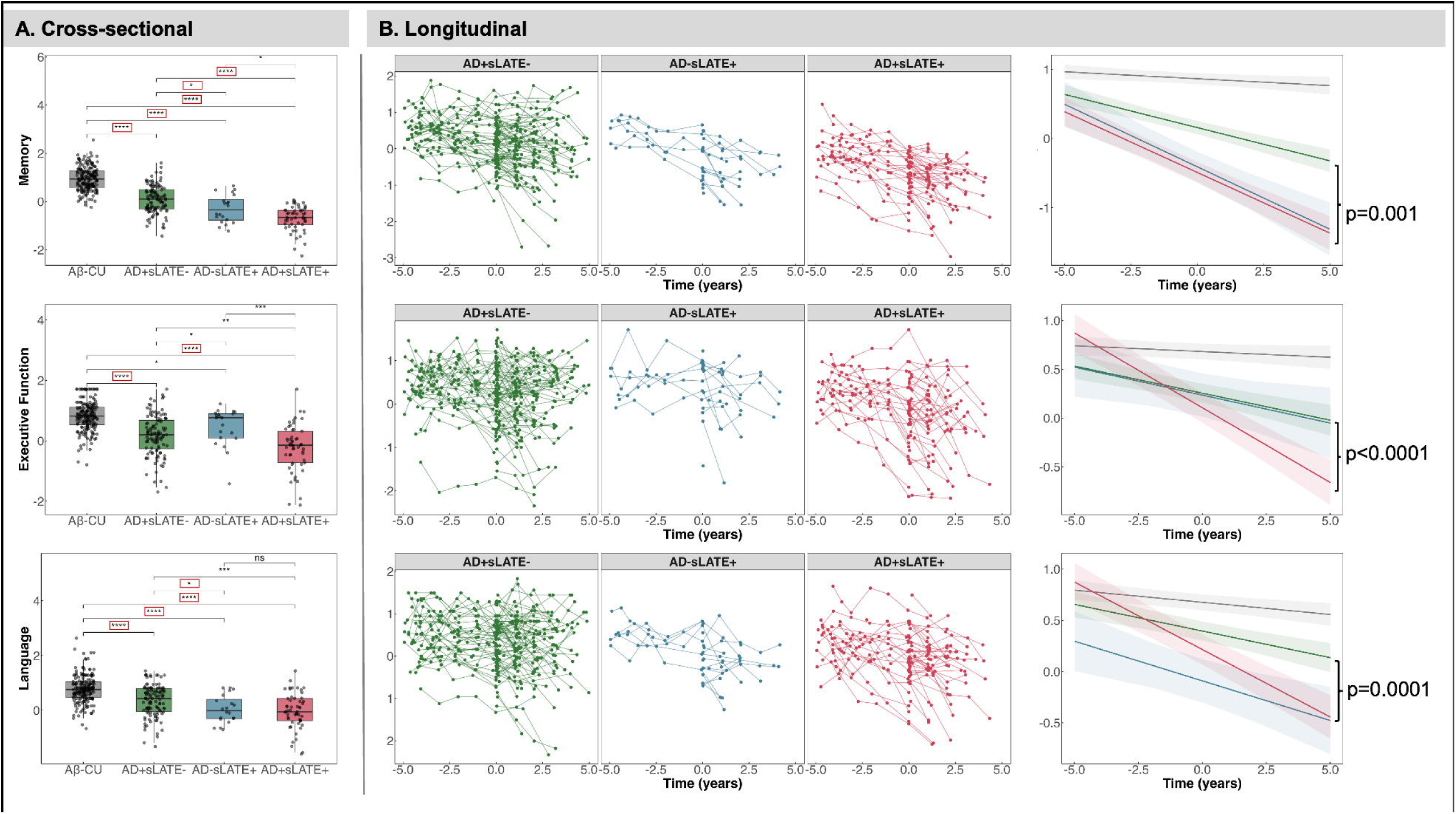
Cross-sectional and longitudinal differences in cognitive domains and trajectories. **A.** Pairwise group differences (*p<0.05, ***p<0.001, ****p<0.0001) at baseline. ** outlined in red reflect significant differences when controlling for global tau max. **B.** Linear mixed-effects model controlling for global tau max and time difference in years across 3 cognitive domains. Domain score ∼ time + group + age at baseline assessment + years of education + Global Tau-MaX + interval between baseline assessment and tau PET + time*group + (1 + time | ID). Model allows for random slopes and intercepts. Year 0 represents the “cross-sectional” timepoint which is anchored to amyloid- and tau-PET imaging. P-values on the right most plot represent significant differences tested between the slopes of AD+sLATE- and AD+sLATE+.

#### 3.4.2. Longitudinal

Longitudinal trajectories across the three cognitive domains were assessed using LME models, adjusting for global tau burden. In the memory domain, AD+sLATE+ had similar decline as AD- sLATE+ but steeper than AD+sLATE-. For executive function, AD+sLATE- and AD-sLATE+ followed a similar trajectory, but AD+sLATE+ showed faster decline. Interestingly, in the language domain, the AD-sLATE+ group started out with lower composite scores than both AD+sLATE+ and AD+sLATE-, but while AD+sLATE+ declined more rapidly over time, AD- sLATE+ showed a slower progression, with rates of decline more comparable to the AD+sLATE- group (Figure 4B).

SILA analyses examining cognitive decline across the three domains, anchored to estimated age of tau onset (see Section 2.4.5), are presented in Figure 5 to illustrate group differences in cognitive trajectories between AD+sLATE- and AD+sLATE+ individuals. All composite scores were scaled by -1 in the SILA models, as SILA associates higher values with greater disease severity; thus, more positive scores indicate greater impairment. Thresholds for cognitive impairment were defined as 1.5 SD above the mean of CU: -0.22 for memory, -0.03 for executive function, and -0.09 for language. When aligning groups along the AD continuum based on estimated age of tau positivity (T+), distinct cognitive trajectories emerged across domains. In the memory domain, individuals with AD+sLATE+ exhibited earlier and more rapid decline than those with AD+sLATE-, with divergence beginning years before tau positivity— suggesting contributions from non-AD pathology first. In contrast, executive function trajectories were relatively similar between groups prior to T+, but the AD+sLATE+ group demonstrated a clear inflection point at tau onset, followed by accelerated decline compared to the AD+sLATE- group. Language performance showed a pattern similar to executive function: while both groups followed a comparable course before T+, the AD+sLATE+ group experienced a steeper decline after crossing the tau positivity threshold. Together, these findings highlight that the presence of LATE co-pathology may drive earlier, pre-tau, memory impairment and more aggressive cognitive deterioration across multiple domains after tau burden becomes substantial.

**Figure 5:**
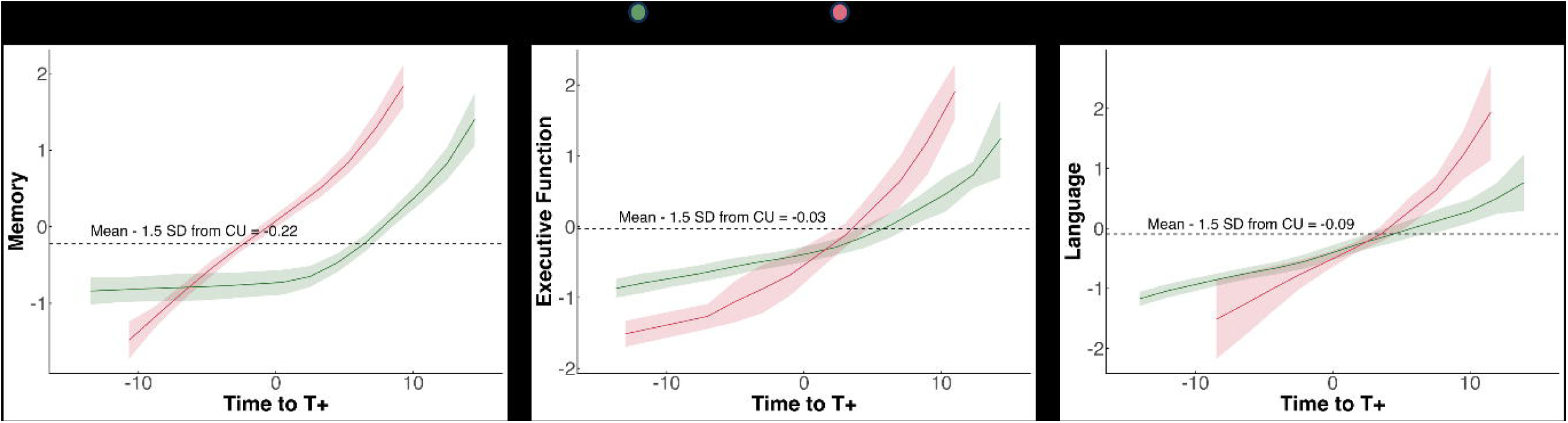
Estimated time to domain-specific cognitive impairment using Sampled Iterative Local Approximation (SILA) modeling. Time to tau positivity (T+) was first estimated using SILA and then anchored to the age at neuropsychological exam. A separate SILA curve was then fit for each group and cognitive domain. The impairment threshold (dashed line) was defined as the mean minus 1.5 standard deviations from CU individuals in ADNI. Composite scores were scaled by -1 so that higher values reflect greater impairment, consistent with SILA’s assumptions. Cognitive trajectories anchored to tau onset age should be interpreted only for groups with tau positivity, i.e., AD+sLATE- and AD+sLATE+.

In addition to aligning trajectories to estimated tau onset (T+), we also examined cognitive trajectories using chronological age at the time of neuropsychological assessment, irrespective of tau status. This approach allowed us to assess domain-specific trajectories across groups without assuming a common pathological anchor point, which is particularly important given that the AD-sLATE+ group does not exhibit tau pathology and therefore lacks a meaningful T+ reference. Notably, under this framework, the AD-sLATE+ group showed more rapid decline similar to AD+sLATE+ in both memory and language domains, further highlighting the LATE-like cognitive profile of the AD+sLATE+ group (Supplementary Figure 2).

### 3.5. Enrichment in autopsy-proven cohort

Among the 79 ADNI participants with autopsy-confirmed neuropathological diagnosis of intermediate/high ADNC, 14 were assigned to the Q1 quartile group and 38 into the Q3-Q4 quartile group based on their adjusted hippocampal volume at baseline MRI. The antemortem interval (AMI), i.e., time between baseline MRI and death, for these 52 participants was 7.2±3.5. Within Q1, 11 of the 14 (78.6%) individuals had autopsy-confirmed diagnosis of LATE-NC (stage 1, 2 or 3), whereas in Q3-Q4, 10 of the 38 (26.3%) individuals had a LATE-NC diagnosis (Figure 6A). These findings underscore that severe hippocampal atrophy seen beyond “typical” AD is associated with concomitant LATE, particularly early in symptomatic disease. Pointwise MTL analyses on baseline MRI revealed expected spatial patterns with autopsy-confirmed AD+LATE+ showing lower thickness in the anterior MTL, particularly the ERC and BA35, compared to autopsy-confirmed AD+LATE- when controlling for Braak stage (Figure 6B). Lastly, the ERC/PHC ratio in the autopsy-confirmed AD+LATE- and AD+LATE+ groups also differ, with AD+LATE+ demonstrating lower ratios, suggesting more anterior atrophy and consistent with our findings in the clinically defined groups (Supplementary Table 1).

**Figure 6:**
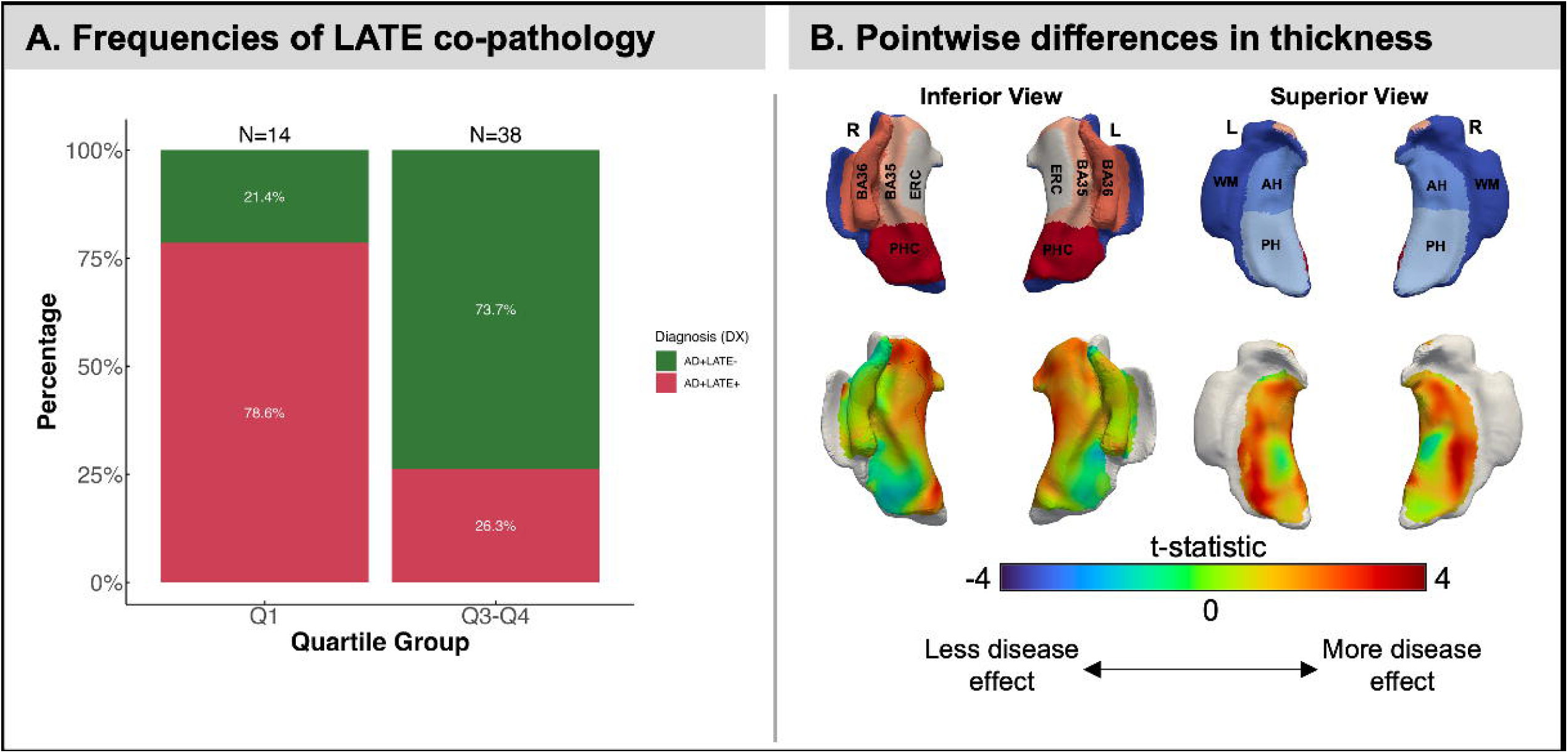
Validation in autopsy cohort using baseline structural MRI. **A.** Frequencies of LATE co-pathology occurrence in low hippocampal volume groups in autopsy confirmed cases. Q1 represents the group with significant hippocampal atrophy and Q3-Q4 represents the group with less severe hippocampal atrophy. **B.** Structural thickness differences in baseline MRI comparing intermediate/high ADNC with and without LATE-NC stage 2 (hippocampal TDP-43), i.e., AD+LATE- > AD+LATE+. Covariates included are age, Braak stage, antemortem interval and field strength (1.5T/3T).

## 4. DISCUSSION

This study demonstrates that a simple, quartile-based cut-off of hippocampal volume effectively identifies a subgroup of cognitively impaired individuals on the AD continuum likely enriched for concomitant LATE and associated with a more aggressive disease course. This stratification approach uses a single MRI-derived metric to group individuals in a way that meaningfully predicts both brain atrophy patterns and cognitive prognosis. The simplicity adds to ease of implementation in both research settings and routine clinical care. The motivation for specifically using hippocampal volume as a probabilistic marker for concomitant LATE emerges from postmortem imaging work in which hippocampal volume is significantly reduced in the setting of LATE with or without concomitant AD.^25^

While other metrics such as the ERC/PHC ratio^21^ and tau-to-neurodegeneration (T/N) mismatch^27,28^ have been proposed as markers of suspected LATE, we chose to focus on whole hippocampal volume for both biological and practical reasons. Hippocampal volume is generally considered a more robust structural measure, particularly in large-scale or heterogeneous datasets. Additionally, T/N mismatch requires tau-PET imaging, which may not be available in many clinical or trial settings, and other PET-based approaches such as FDG-PET^22–24^ face similar constraints, limiting their broad applicability. In contrast, hippocampal volume can be derived from a single structural MRI scan, increasing its potential for integration into both research pipelines and clinical workflows.

Using this cut-off, we found that individuals in the lowest hippocampal volume quartile (Q1) displayed significantly different imaging and cognitive profiles from those in the higher quartiles. The group identified as AD+sLATE+—characterized by low hippocampal volume in the context of biomarker confirmed AD—displayed pronounced thinning in the anterior MTL, including the ERC and BA35—regions previously implicated in TDP-43 pathology. In addition to anterior MTL thinning, the AD+sLATE+ group also exhibited reduced amygdala volumes, consistent with recent autopsy studies identifying lower amygdala volumes in individuals with LATE-NC.^11,46,47^ This finding provides further support for LATE involvement, as the amygdala is known to be an early and vulnerable site for TDP-43 pathology. The ERC/PHC ratio, a metric previously demonstrated to discriminate between cases of AD with versus without TDP-43 in a postmortem series^21^, was also lower in the AD+sLATE+ group. Notably, the AD-sLATE+ group had a similar reduced ratio compared to the AD+sLATE- group. In addition, both AD+sLATE+ and AD-sLATE+ groups exhibited higher asymmetry indices, supporting literature linking LATE with asymmetric atrophy patterns.

Extending beyond the MTL, the AD+sLATE+ group displayed more widespread cortical thinning, particularly in regions associated with LATE, such as medial-anterior temporal polar and orbitofrontal cortex and anterior extrahippocampal region. Notably, the spatial pattern of atrophy observed here closely mirrors regional vulnerability reported in a recent ex vivo MRI study examining imaging correlates across neuropathologically defined LATE-NC stages.^48^ Similar patterns have also been demonstrated in structural MRI and FDG-PET studies of LATE-NC, and converge with the known distribution of TDP-43 pathology described in postmortem investigations.^23,49,50^ This consistency across imaging modalities and pathological studies reinforces the biological plausibility of the phenotype observed here as reflecting suspected TDP-43 co-pathology. Longitudinally, AD+sLATE+ exhibited faster cortical thinning than either AD+sLATE- or AD-sLATE+, particularly in anterior MTL and temporal polar regions, even after adjusting for tau burden—reinforcing the likely role of co-pathology rather than AD alone in driving this difference.

The cognitive differences between these groups largely mirrored expectation based on the imaging. AD+sLATE+ individuals had lower memory and language performance at baseline and declined more rapidly over time. Of particular note, memory decline in this group appeared to precede estimated tau positivity. As cognitive decline generally does not appear prior to tau burden measurable with PET imaging, this result suggests that LATE, or another non-AD pathology is likely to contribute to these memory changes prior to manifest clinical AD in at least a subset of these patients. For language and executive domains, the AD+sLATE+ group followed a similar trajectory to AD+sLATE- prior to tau positivity but had an accelerated drop-off after reaching tau thresholds—indicating a potential synergy between AD and TDP-43 pathologies in driving decline. This finding is also consistent with LATE having its earliest and most prominent effect on memory.^15^

Importantly, we further validated the quartile-based hippocampal volume threshold in an autopsy-confirmed cohort. Among individuals who fell into the lowest hippocampal volume quartile at their first MRI in the ADNI study nearly 80% had confirmed AD+LATE+. Alternatively, only about 20% with ADNC at autopsy without LATE met this threshold at their first scan. As symptomatic patients in ADNI are enrolled at either the MCI or mild dementia stage, and some were even cognitively unimpaired in this autopsy group, this supports the notion that severe atrophy at early disease stages is enriched in those with concomitant LATE. As this population is also the target population for recently FDA-approved anti-amyloid therapies, the threshold developed here may be a useful tool for stratifying patients and determining whether suspected LATE co-pathology influences outcomes.

While the quartile-based hippocampal volume cut-off used here clearly enriches in individuals with concomitant LATE, it does not definitively distinguish the presence of TDP-43 co-pathology from other potential drivers of hippocampal atrophy. For example, non-AD pathologies, such as argyrophilic grain disease (AGD), frontotemporal lobar degeneration (FTLD), including other TDP-43 pathologies, and cerebrovascular disease. Alternatively, the so-called limbic variant of AD also may present with more severe MTL atrophy. Incorporation of the cut-off described here with other clinical features from the recently proposed LATE criteria may result in further specificity. In particular, disproportionate hippocampal atrophy relative to MTL tau burden measured by tau-PET may reduce the likelihood of limbic variant AD. Moreover, the observation of marked atrophy even in cases without confirmed LATE highlights the broader complexity of medial temporal lobe vulnerability in aging and neurodegeneration. These limitations underscore the ongoing need for validated in vivo biomarkers specific to TDP-43 to improve pathological specificity.

This study has several limitations. While we validated our stratification approach using autopsy-confirmed cases, the cohort size was modest, and the cut-off was derived cross-sectionally, which may not fully capture the longitudinal progression of LATE-related neurodegeneration.

Additionally, this work was conducted in ADNI—a highly selected research cohort—and findings may not generalize to more diverse or community-based populations, where the prevalence and manifestation of co-pathology may differ.

In the absence of molecular biomarkers for TDP-43, structural imaging measures offer a probabilistic link to LATE, even in the presence of AD pathology. As indicators of downstream neurodegeneration, these measures provide not only a signal of potential co-pathology but also insight into disease stage, based on the pattern and severity of atrophy. This added dimension can support clinical decision-making, enhance patient stratification, and inform therapeutic targeting—particularly in trials where identifying individuals with mixed pathology is essential.

The core contribution of this work lies in demonstrating that a simple, normative MRI-based threshold—readily implementable across clinical and research settings—can yield biologically and clinically meaningful groupings. This approach offers a scalable strategy for enriching clinical trial cohorts, flagging high-risk individuals in practice, and improving our understanding of the complex interplay between co-pathologies in neurodegenerative disease. Depending on the therapeutic objective, this stratification method could be used to either selectively exclude individuals with suspected mixed pathology in trials seeking pure-AD cohorts or intentionally include them when targeting the broader biological complexity of AD with co-existing LATE.

## Data Availability

All data produced in the present study are available upon reasonable request to the authors.

## Notes

### Competing Interest Statement

N.S.M., X.L., C.A.B., N.S., E.M., P.A.C., J.G., P.A.Y., and S.R.D., declare no competing interests. Long Xie is a paid employee of Siemens Healthineers. David Wolk has served as a paid consultant for Eli Lilly and Beckman Coulter. He has also served on the DSMB for Functional Neuromodulation and GSK. He has received research support paid to his institution by Biogen.

### Funding Statement

This work was supported by grants from the National Institutes of Health (P30-AG072979, RF1-AG069474, R01-AG056014, R01-AG072796), and Alzheimer's Association and Fred Barbara Erb Foundation (AACSF-23-1152241). Data collection and sharing for the Alzheimer's Disease Neuroimaging Initiative (ADNI) is funded by the National Institute on Aging (National Institutes of Health Grant U19 AG024904).

### Author Declarations

Details about ADNI informed consent procedures are available from www.adni-info.org.

