## Supplemental Figures+Tables for "Enrichment of patients with concomitant LATE on the Alzheimer’s disease continuum using hippocampal volume"

**Supplementary Figure 1: Cohort Flowchart.**

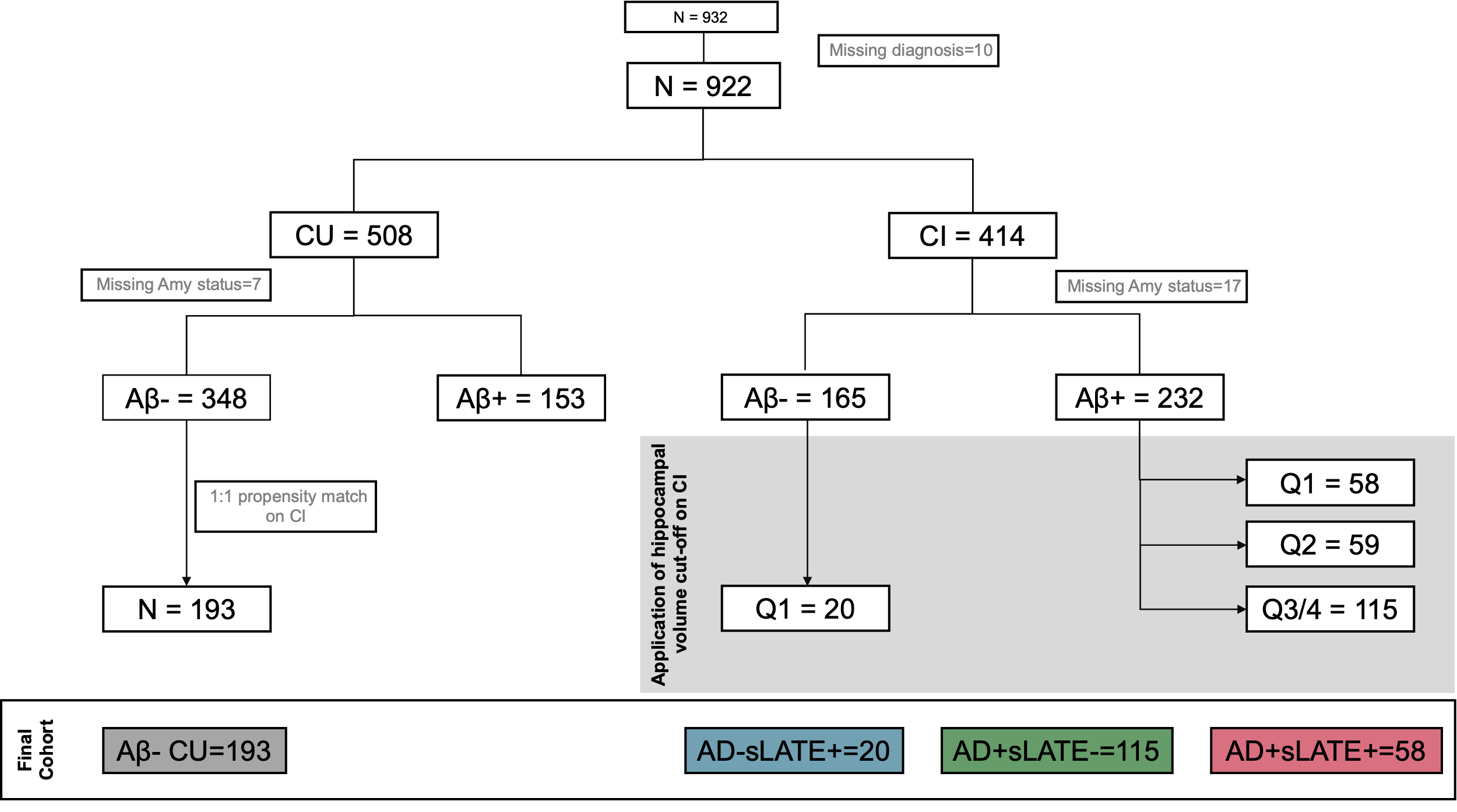

**Supplementary Figure 2: Estimated progression to domain-specific cognitive impairment across chronological age using SILA modeling.**

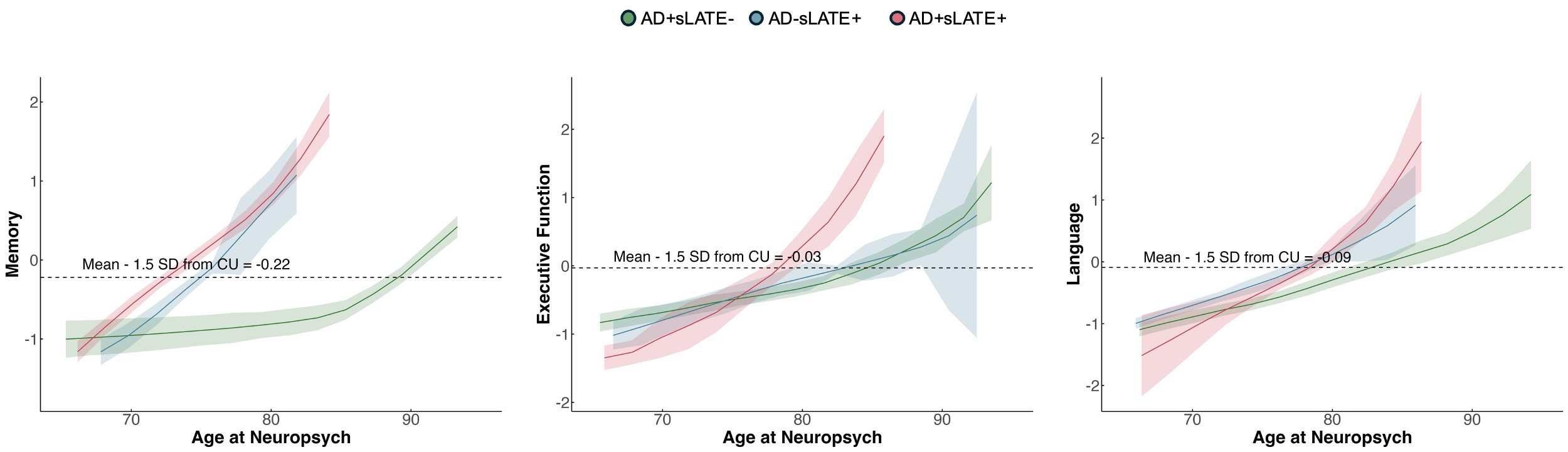

**Supplementary Table 1: Demographics and Imaging Features in Autopsy Cohort.**

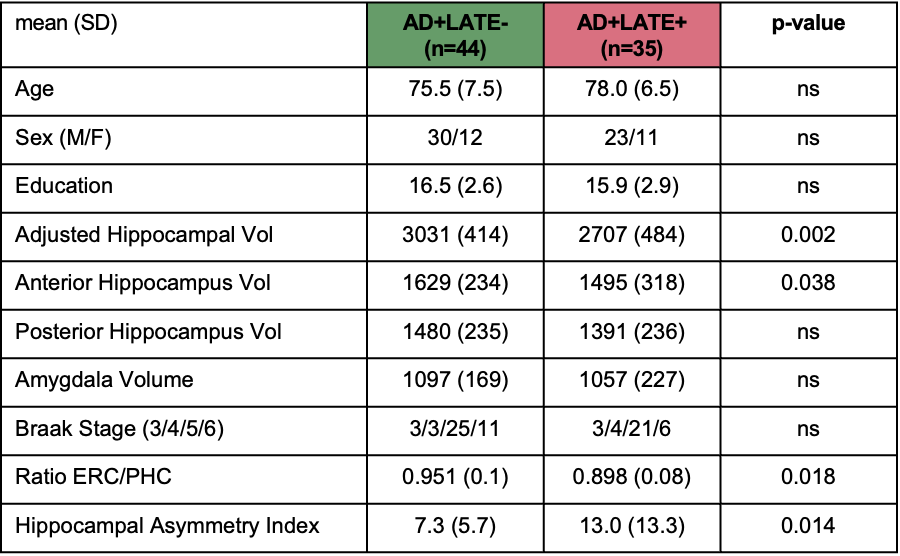
